# Healthcare professionals’ perspectives on a multilevel cardiovascular risk management intervention (PROSPERA programme)

**DOI:** 10.64898/2026.06.08.26355169

**Authors:** Vera A. M. C. Bongaerts, Laurens C. van Gestel, Petra G. van Peet, Marie-Lise S. Vuijk, Steven H. J. Hageman, Jannick A. N. Dorresteijn, Tobias N. Bonten, Mattijs E. Numans, Hendrikus J. A. van Os, Rimke C. Vos

## Abstract

**Background:** Two-thirds of Dutch cardiovascular risk management (CVRM) for patients at risk of cardiovascular disease is delivered in primary care practices. While individual risk scores are increasingly used during consultation, a population-level structure for risk-based patient outreach is not currently available. We therefore developed the PROSPERA programme, a multilevel intervention comprising population-level risk stratification and individual-level support tools.

**Aim:** To assess anticipated and experienced barriers and facilitators among healthcare professionals (HCPs) to inform implementation in primary care.

**Methods:** We conducted four focus groups and six interviews with nine primary care HCPs to explore anticipated and experienced barriers and facilitators. Inductive codes were thematically analysed and assigned to corresponding domains of the Theoretical Domains Framework (TDF) and the related Capability, Opportunity, Motivation model of Behaviour.

**Results:** Barriers and facilitators were identified in 11 TDF domains. Population-level barriers included altered professional roles and limitations in technological infrastructure. Individual-level barriers were limited skills in interpreting risk calculations and difficulty integrating tools into clinical routine. Facilitators were related to beliefs on the importance of providing proactive care (population level), the use of U-Prevent for risk communication (individual level) and positive patient responses to the Lifestylecheck questionnaire (individual level).

**Conclusion:** Addressing barriers and facilitators identified at both the population and individual levels can support implementation of the PROSPERA programme. Opportunities exist in education and training of HCPs in risk communication, as well as support in restructuring the physical and digital environment.

## INTRODUCTION

Cardiovascular disease (CVD) is the most important cause of mortality worldwide and its prevalence is rising (1, 2). In the Netherlands, about two third of cardiovascular risk management (CVRM) is delivered in primary care via integrated care programs (3). These are programs coordinated by the general practitioner (GP) and characterised by interdisciplinary collaboration of (para)medical disciplines to ensure continuity of care for patients with chronic conditions. Since the introduction of these programs in the Netherlands, cardiovascular prevention is increasingly the responsibility of practice nurses working under GP supervision (4).

HCPs in primary care follow CVRM guidelines, which provide the GP and the practice nurse with protocolized standards (5, 6). Although guidelines advise calculating individual risk scores for patients during consultation, a population risk-stratification structure to prioritize CVRM is not currently in place. In addition, discussing individual risk scores and using treatment modelling to prioritise personalised treatment goals together with the patient is not yet universal in daily practice. As a result, CVRM tends to follow a relatively uniform approach, with limited differentiation across patient subgroups. Available resources, including the attention and capacity of HCPs, are therefore not always distributed equitably across this heterogeneous population (7, 8).

The PROSPERA programme, using a population health management approach to proactive care, was developed to provide risk-dependent CVD prevention in an era of limited resources (9). This multilevel complex intervention is aimed at the primary care population with increased cardiovascular risk, including apparently healthy patients, patients with established CVD and patients with diabetes mellitus (DM). The PROSPERA programme comprises population-level risk stratification and three individual-level components: a lifestyle questionnaire (Lifestylecheck), an application for interactive cardiovascular risk calculations (U-Prevent) and optional remote patient monitoring tools (the Box). Effectiveness of the programme will be examined in a separate future cluster randomized controlled trial (10).

However, demonstrating the effectiveness of a complex intervention does not necessarily result in its uptake in clinical practice (11, 12). Previous studies examining the research-to-practice pipeline found that only a small proportion of health research (4% to 14%) is translated into sustainable clinical practice (13-16), and this number may be even lower for complex interventions. Since the return on investment for society lies in sustainable adoption, research focusing on early stage barriers and facilitators is crucial to the selection of effective implementation strategies (17-19).

Therefore, the current study assesses anticipated and experienced barriers and facilitators among HCPs to inform implementation of the PROSPERA programme in primary care.

## METHODS

### Design

A qualitative study was conducted using focus groups and semi-structured interviews with HCPs. Topics were consecutive and discussed in a sequential order. For theoretical embedding we used the validated Theoretical Domain Framework (TDF) and the related Capability, Opportunity, Motivation model of Behaviour (COM-B), as described by Michie et al. (17, 18, 20). Both models are widely used in implementation research to assess determinants of HCP behaviour and provide a basis for the selection of appropriate implementation strategies (18, 19).

### Population

Since the Extramural Leiden University Medical Center Academic Network (ELAN) served as the data source for the PROSPERA programme, only HCPs from primary care centres affiliated with ELAN were invited to participate. GPs and practice nurses were eligible if they worked in areas serving patients with various socioeconomic backgrounds. Practice nurses were eligible if involved in CVRM. HCPs were subsequently purposively sampled based on their use of different types of electronic health records (EHRs) and their availability to participate in all three focus groups, given the sequential nature of the topics discussed across the sessions. Participants were asked to provide written or digital informed consent.

### PROSPERA programme

Development of the PROSPERA programme followed the Medical Research Council (MRC) framework for developing and evaluating complex interventions (21). The version of the PROSPERA programme in the current study comprises a population health management approach that applies proactive population-level risk stratification to categorize patients at risk of CVD into panels, as well as three individual-level components: i) a pre-consultation Lifestylecheck questionnaire focused on the motivation of the patient for lifestyle change, ii) the U-Prevent clinical decision support tool for individual risk communication and iii) the Box for remote patient monitoring, containing a blood pressure monitor, a weighing scale and an activity meter. Figure 1 shows a schematic visualisation.

**Figure 1.**
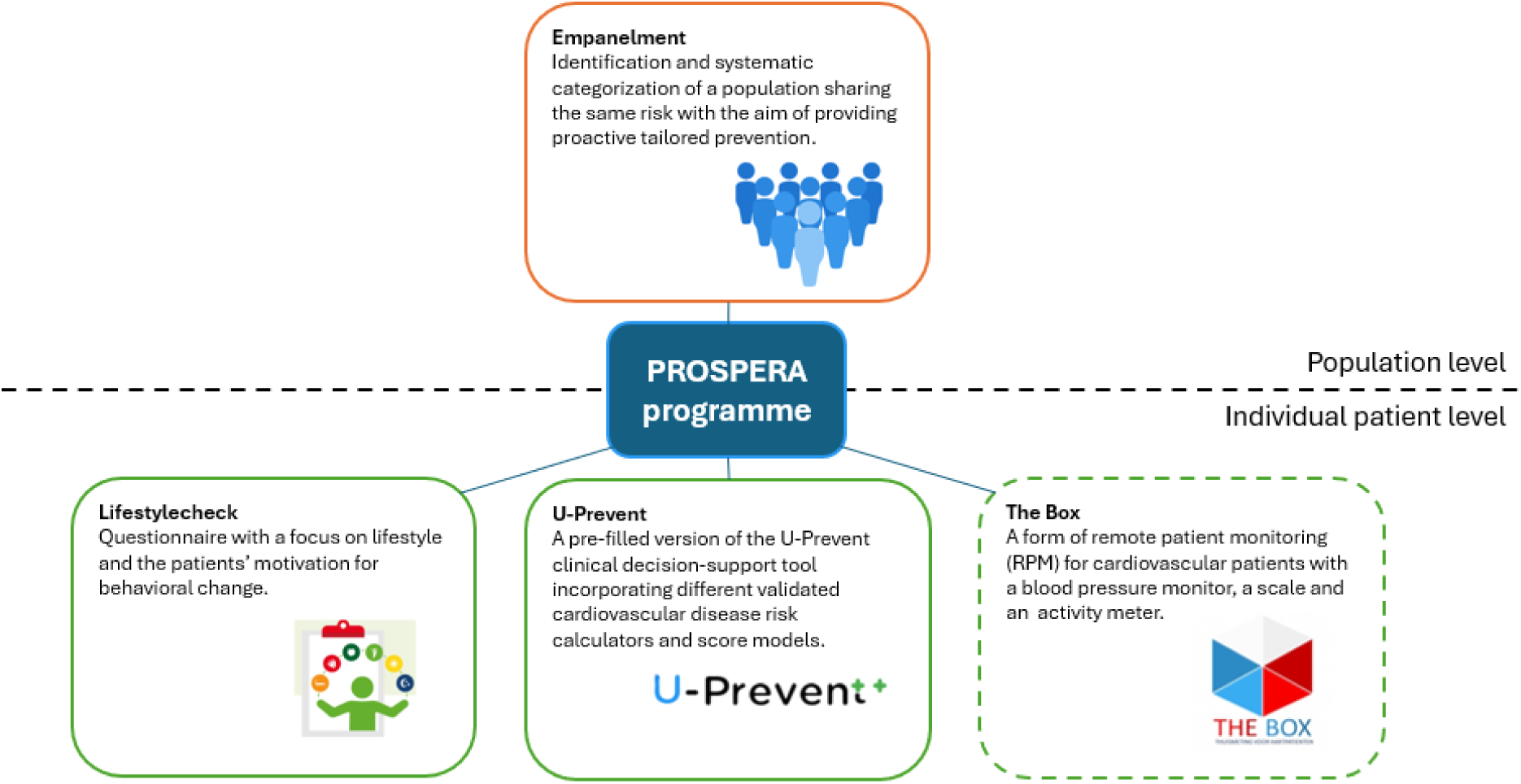
Components of the PROSPERA programme.

### Target behaviours

Implementation of the PROSPERA programme involves a change in workflow at both the population and individual levels and as such requires HCP behavioural change. Therefore, two target behaviours were specified following the first stage from the Behavioural Change Wheel (17):

1. **Providing proactive care at the population level** The HCP prioritises patients for proactive cardiovascular prevention. To support this, the researcher provides the HCP with a list of patients at risk of CVD stratified to three panels: normal, high, extra high panel. This stratification was based on whether the patient required primary or secondary prevention and whether the patient had achieved his or her preventive CVD treatment goals. Further details on panel creation are available in the PROSPERA protocol manuscript (10). The HCP can use this list, together with their professional judgment, to prioritise patients for outreach and to determine consultation follow-up frequency.
2. **Using support tools at the individual level** The HCP uses the Lifestylecheck, U-Prevent and the Box (optional) at the individual level to prioritise treatment and treatment targets. This means that the HCP needs to interpret the results prior, during or after consultation with the patient and that the HCP and patient engage in shared decision making concerning the treatment plan.

### Data collection

The topic lists for the focus groups and interviews were based on the 14 TDF domains (see supplement *Topic List*). The focus groups were designed and conducted as a sequence to address barriers and facilitators at all operational stages of the PROSPERA programme. Focus groups topics and interviews are shown in figure 2.

**Figure 2.**
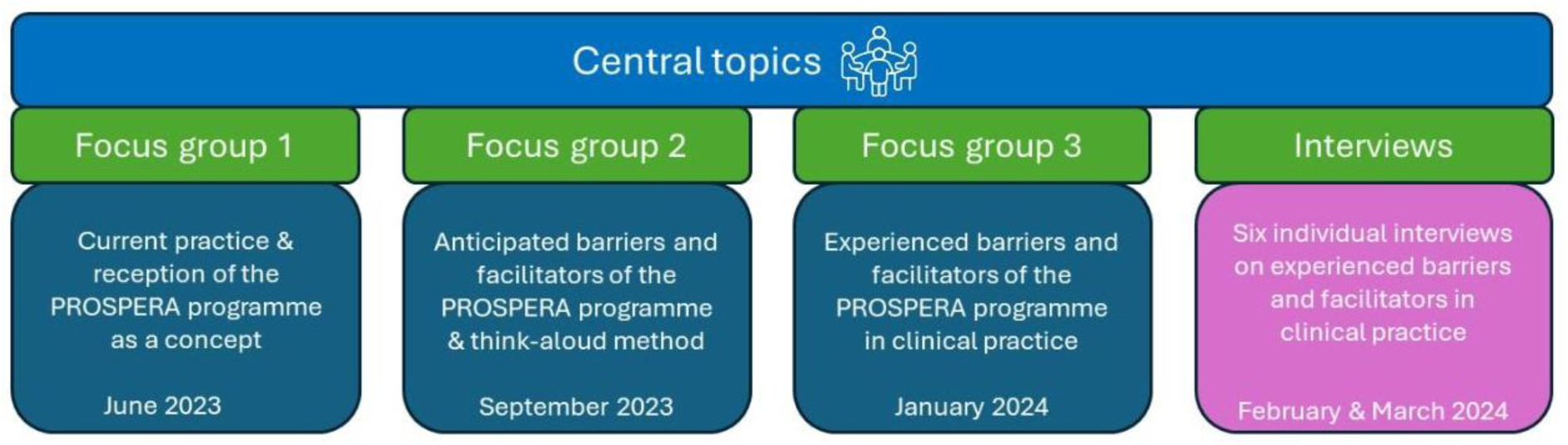
Central topics of focus group sessions and interviews.

The first in-person focus group took place in June 2023 and concentrated on barriers and facilitators experienced in current integrated care programs for patients at increased risk of CVD. For practical reasons and to broaden the scope, a digital complementary focus group on the same topic was held with two additional HCPs who were unable to attend the in-person session. The second in-person focus group was held in September 2023 with the same cohort of HCPs and contained a pilot version of the PROSPERA programme. It focused on *anticipated* barriers and facilitators when using the PROSPERA programme, and included a classic think-aloud method as developed by Ericsson and Simon (1993) to assess the dashboard function and the ICT infrastructure of the PROSPERA programme (22). Following the second focus group, a pilot version of the PROSPERA programme was introduced in 3 affiliated general practices of participating HCPs in The Hague region. The third and final in-person focus group was held in January 2024 and concentrated on *experienced* barriers and facilitators when using the PROSPERA programme in clinical practice. As the HCPs continued working with the intervention for another month, additional semi-structured individual interviews on experienced barriers and facilitators were conducted with the same cohort of HCPs in February and March 2024 to complement previously generated data. All interviews took place on location at the respective general practices. Focus groups were scheduled to last between 90-120 minutes; interviews 30-60 minutes.

### Data analysis

All focus groups and individual interviews were audio-recorded and transcribed verbatim. Two researchers (VB, MV) independently coded the transcripts using *Atlas.ti 23*. The inductive codes were thematically analysed, classified as a barrier or facilitator, and assigned to corresponding TDF domains and COM-B constructs for one of the two specified target behaviours. The established codes were compared and discussed by two researchers (VB, MV). If necessary, a third researcher (RV or LG) was included until consensus was reached.

### Positionality

The research team represented diverse professional backgrounds and varying levels of experience. VB is a GP trainee with prior experience in treating CVD in general practice. MV is a medical student with limited experience in the field of CVD. LG is a behavioural scientist with a focus on healthcare professionals’ behaviour, and RV a public health scientist specialised in implementation science and cardiometabolic health. None of the researchers had personal experience with the study topic and there were no prior relationships with the participants.

## RESULTS

Nearly all participants were female, with work experience ranging from 3 to 30 years as a GP (n=5) or practice nurse (n=4). Barriers and facilitators were identified in 11 out of 14 TDF domains and organized below according to the COM-B for each specified target behaviour.

### Providing proactive care at a population level

#### Capability

In current CVRM practice (i.e., in the absence of a data-driven tool), HCPs experienced difficulty scheduling patients for prioritised outreach. They expressed the wish for a tool to support identification of their top 5 patients at risk of CVD in need of intensive follow-up, noting that they were often unable to recall patients by memory due to limited familiarity with their patient population (*memory attention and decision processes*) (#Q0).

> ***Q0.*** *‘Data will support you in certain decisions because you no longer know the patient that well.’*

#### Opportunity

As regards proactive care, HCPs anticipated and experienced several barriers and facilitators within the *environmental context and resources* domain. They anticipated that patient categorisation into panels (normal, high, or extra high) would help prioritise whom to target for proactive preventive care. However, a few professionals experienced time constraints, since the panel list displayed all patients rather than only those assigned to them. Prioritising patients therefore sometimes resulted in a cumbersome process (#Q3). Another experienced barrier was the PROSPERA panels’ limited accuracy capturing all eligible patients at risk of CVD (#Q16).

> ***Q16****. ‘It seemed to me that the list wasn’t as accurate as it should have been: dates of birth, certain selections that didn’t correspond with chronic CVRM care…’*

HCPs indicated two possible causes. First, as real-time data were not yet available, the frequency of data extraction from EHRs was limited and newly enrolled patients were not enlisted instantaneously. Second, poor EHR documentation might have resulted in misclassification of patients in integrated care programs. A final experienced barrier to proactive care in the *environmental context and resources* domain was the study’s elaborate and time-consuming informed consent procedure (#Q5).

Although HCPs anticipated that the PROSPERA panels alone would not be sufficiently conclusive to determine recall- or follow-up frequencies (#Q21), they nonetheless experienced it as valuable and complementary to their own knowledge and decision making process.

> ***Q21****. ‘That is what I find interesting about the first step: creating the panels relies more on data rather than on intuition. And then of course other factors play a role: level of education etc. Clearly, not everything can be found in data. That’s why it’s tricky.’*

GPs specifically anticipated that the PROSPERA panels would support practice nurses to work more autonomously. This was endorsed by practice nurses, who anticipated that the panels would support their strategic planning of follow-up consultations.

#### Motivation

Despite the experienced limitations of the existing invitation system, HCPs indicated low confidence and uneasiness when anticipating replacement of the present structure with a new proactive approach (*beliefs about consequences*). Nevertheless, one anticipated facilitator for engaging in proactive care was to explore the possibility of identifying patients in need of immediate or intensive follow-up (#Q9). Similarly, HCPs indicated equal interest in identifying patients needing fewer follow-up consultations (*goals*) (#Q4, Q11).

> ***Q11****. ‘I’m also a big proponent of identifying people who don’t require consultation time. I’m talking about the people with a CVRM risk profile who are on a statin and take their blood pressure medication, and who’ve been stable for years and can self-manage. To be honest I’d almost want to exclude them from check-ups*.

GPs anticipated a barrier, however, in the self-confidence and self-determination of practice nurses in deciding when to deviate from routine follow-up or recall frequencies independently of GPs *(beliefs about capabilities)* (#Q23).

> ***Q23****. ‘Look, GPs are willing to disregard their guidelines or to follow a gut feeling, but practice nurses are much less trained in doing that. They work within the guidelines and are used to ticking all the boxes. And they really aren’t willing to [abandon that]. So, for a practice nurse to make a decision like that… you need some sort of guidance*.’

Some HCPs, in particular practice nurses, were concerned about decreasing follow-up frequencies or replacing in-person consultations with remote patient monitoring (for patients in the normal panel). They were accustomed to seeing the patient annually and felt reluctant to alter this routine, feeling that it required a loosening of control and adopting a different professional attitude. They also felt that remote patient monitoring or telephone consultations could never fully replace a physical comprehensive evaluation and will consequently alter the professional-patient relationship (*professional role and identity*).

HCPs were ambiguous concerning who bore responsibility for population risk stratification. All preferred a concise, readily-interpretable overview with clear directives on who to invite for consultation, stating that population risk stratification itself, or automation and optimization of the procedure, should be carried out and supported by regional care groups (#Q10) (*professional role and identity)*.

### Using support tools at the individual level

#### Capability

In the domain *behavioural regulation*, a facilitator was that all HCPs actively planned when and how to discuss the Lifestylecheck questionnaire and the pre-filled version of U-Prevent during the consultation (#Q18). They often paid explicit attention to the patient’s ‘homework’ and began a consultation with the patient’s completed pre-consultation Lifestylecheck responses (#Q12).

> ***Q12.*** *‘I started with the Lifestylecheck because people sometimes prefer to begin with something simple instead of something that’s immediately in-your-face. And I also feel that if people do something for you, even if it’s just filling in a sheet of A4, then you should give it the attention it deserves.’*

However, HCPs already using the Lifestylecheck found it difficult to alter routine and consequently continued to answer questions together with their patients during consultation (*behavioural regulation*). A few HCPs unfamiliar with the Lifestylecheck mentioned being already so overwhelmed by the prefilled U-Prevent application (also new), that they sometimes forgot the Lifestylecheck questionnaire (#Q13) *(memory, attention and decision processes*). While overall understanding of U-Prevent was anticipated to be good, its use in clinical practice led to questions about the treatment modellers and functional modalities of the risk calculators, especially if a patient did not seem to clearly fit either of the calculators (*knowledge*).

#### Opportunity

HCPs mentioned that the cardiovascular patient population generally showed low literacy, which they experienced as a barrier for providing the Lifestylecheck prior to the consultation as intended (#Q14) *(environmental context and resources)*. In addition, HCPs also anticipated that the Box would only be feasible for specific patients capable of self-management (#Q6) *(environmental context and resources)*.

> ***Q6.*** *‘That’s what we’re talking about. We’re talking about the KLM model. Who’s a captain? Who’s a flight attendant? Who’s a passenger? And who’s a passenger with luggage?’*

In relation to U-Prevent, practice nurses mentioned the importance of preparation time and proper instruction for skill development in risk communication (*environmental context and resources*) (#Q34).

#### Motivation

Patient completion of the Lifestylecheck questionnaire before the consultation was experienced as a facilitator by half of HCPs, as this activated patients to examine their wellbeing and motivation for lifestyle change beforehand (#Q2).

> ***Q2***. ‘*Completing the Lifestylecheck pre-consultation shows that people have already thought about ‘What does my life look like across the board? What can I change? What effect might that have?’ I noticed that, for those few people, it does have an impact - it gets their thought processes moving. And, as I’ve said before, it was a great way to start a conversation.*’

HCPs also anticipated that visualisations of calculated risk scores and treatment effects in U-Prevent would be a facilitator. They believed that the concept of risk would be easier to explain through the coloured bars and graphs *(beliefs about capabilities*) (#Q22).

> ***Q22.*** *‘People who are less educated often demonstrate a strong visual awareness. That means I think, that regardless of your perspective, it’s beneficial to use U-Prevent to show and explain the concept of risk.’*

However, in the experience of some HCPs, visualization did not always translate to a deeper understanding (#Q7). One GP anticipated that the U-Prevent/Lifestylecheck combination would activate and enhance patient self-management (*beliefs about consequences*) (#Q8). However, GPs anticipated a barrier in the interpretation of calculated risk scores and treatment benefits by practice nurses, since these skills were not part of their vocational training (*beliefs about capabilities)* (#Q1). After using U-Prevent in clinical practice, both practice nurses and GPs confirmed their earlier expectations: they felt it was relatively intuitive but expressed need for additional training in risk communication *(skills).* Overall, HCPs described a feeling of nervousness and excitement upon first use of the Lifestylecheck and U-Prevent (*emotion*) (#Q17). This was not considered a barrier, just a natural reaction to a new approach.

Most HCPs mentioned they had no intention of using the Box according to the original PROSPERA design (*intentions*). They supported temporary loans of blood pressure monitors for periodic assessment but opposed continuous use as they anticipated a need for constant vigilance over high readings.

**Table 1.**
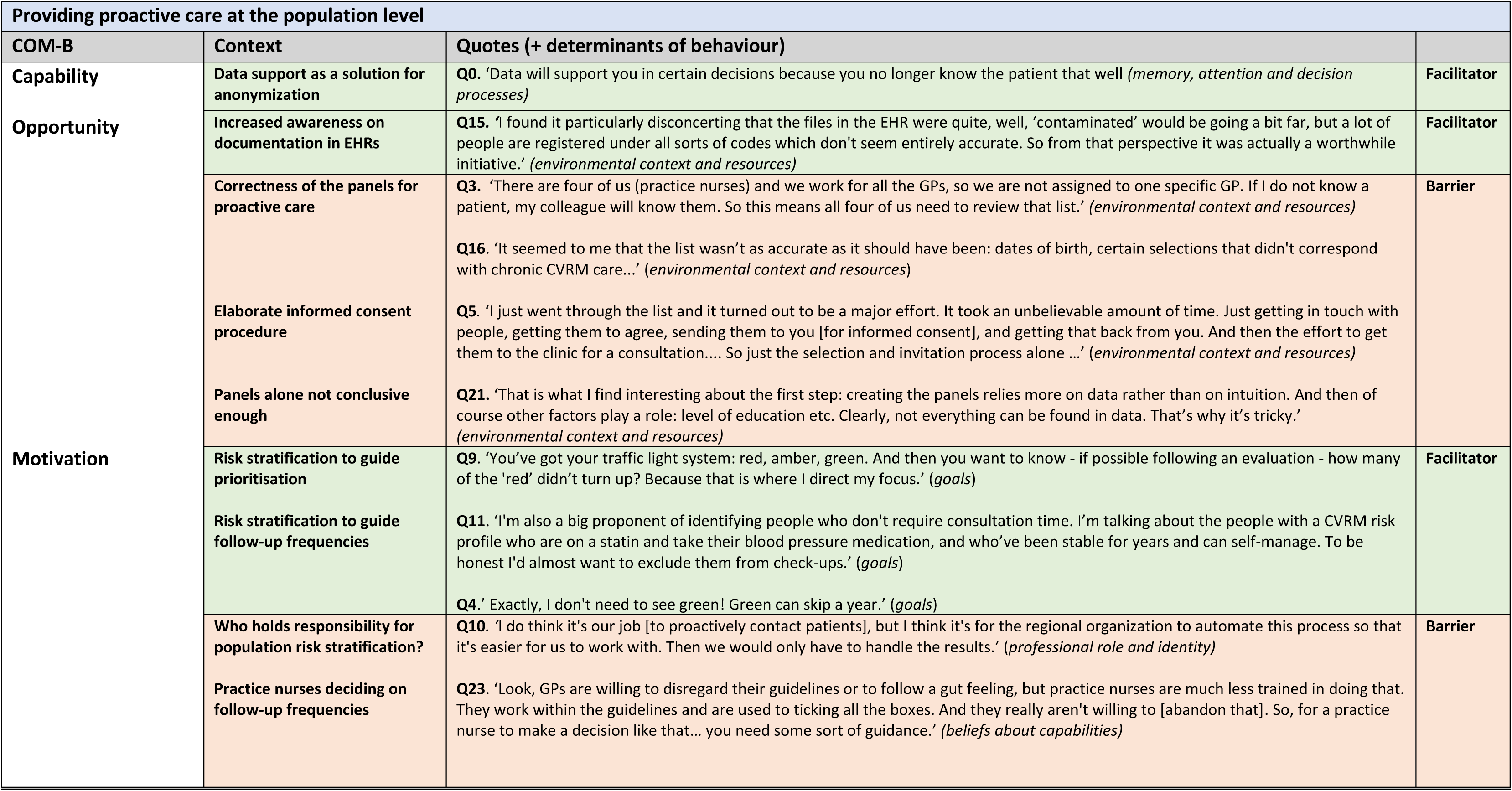
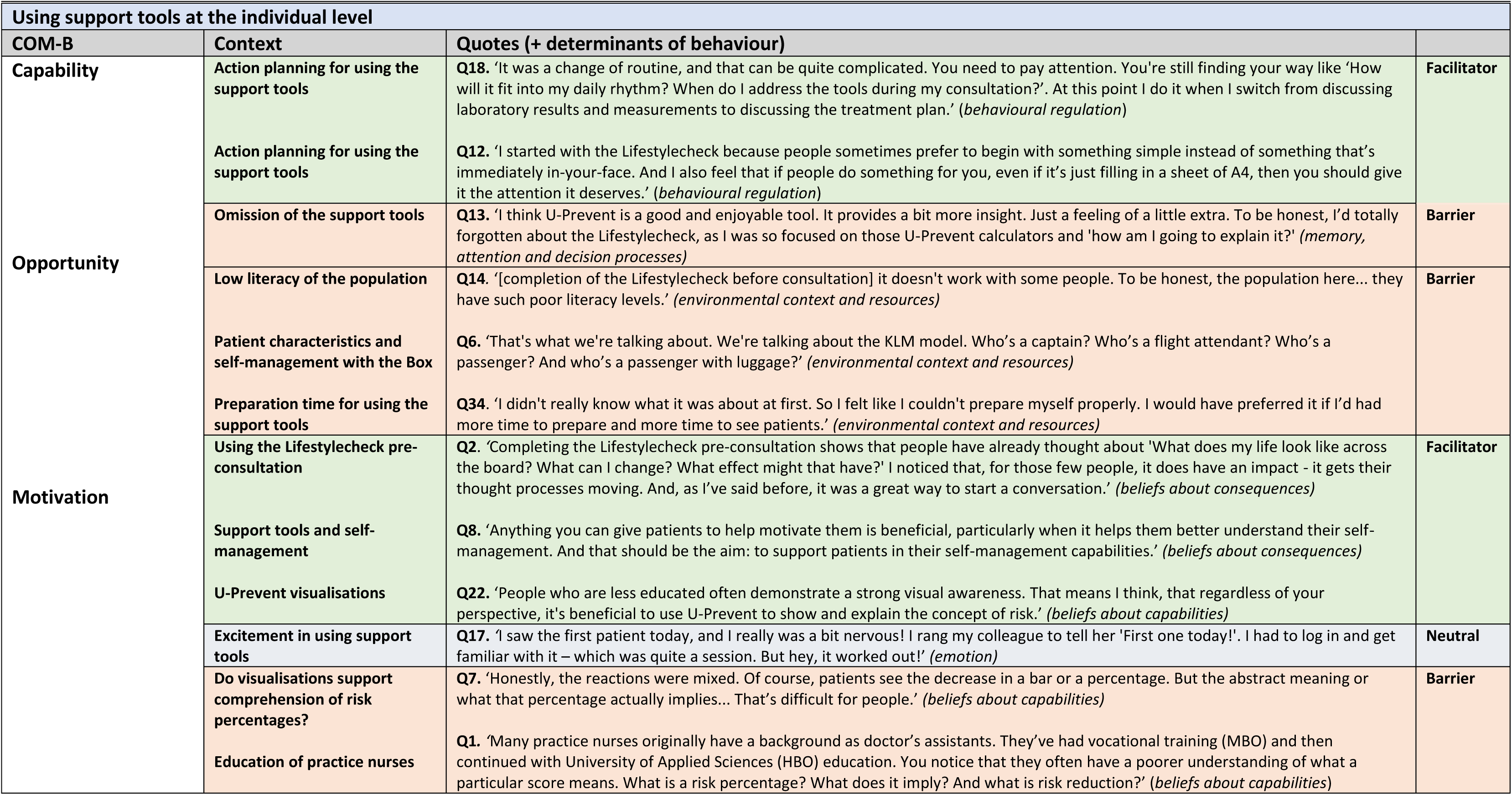
Barriers and facilitators of using the PROSPERA programme per specified target behaviour.

## DISCUSSION

### Summary

For both population- and individual-level target behaviours, we identified barriers and facilitators that inform implementation of the PROSPERA programme in primary care. Regarding ‘providing proactive care at the population level’, the main barriers related to creating panels and elaborate study procedures (all *environmental context and resources*). Other barriers concerned uncertainty about the ability of practice nurses to manage follow-up frequency independently of GPs (*beliefs about capabilities*), and the possible negative impact of reduced consultation frequency on the professional-patient relationship (*professional role and identity*). A main facilitator was the motivation of HCPs to prioritise patients according to care needs (*goals*). For ‘using support tools at the individual level’, experienced barriers were breaking habits during clinical routine (*behavioural regulation*), unintentional omission of the Lifestylecheck (*memory, attention and decision processes*), low patient literacy (*environmental context and resources*) and uncertainty about practice nurse skills in interpreting U-Prevent risk scores (*beliefs about capabilities*). Main facilitators were positive patient responses to the Lifestylecheck (*beliefs about consequences*) and the contribution of graphs and visuals in the clinical decision tools to support risk communication (*beliefs about capabilities*).

### Strengths and limitations

A strength of this pilot study was that implementation of the PROSPERA programme was considered a priority from the start (17). We explored both anticipated and experienced barriers and facilitators at an early phase, attentive to potential conflicts or alignment of the desired HCP target behaviours with behaviour in current clinical practice. Another strength was the decision to conduct consecutive focus groups with the same HCP cohort, ensuring uniform understanding and an equal exchange of experiences. A further strength was HCP sampling (‘the target adopters’) from general practices in relatively low-income areas, serving patients from a broad range of socioeconomic backgrounds.

Since this perspective is often difficult to capture, this is considered a key asset. As HCPs are the main stakeholders, successful adoption depends on their receptiveness and willingness for behaviour change. A final strength therefore was the use of the TDF to provide structure, focusing explicitly on GP and practice nurse behaviour when using the PROSPERA programme (17, 18).

However, focus groups and interviews also have limitations, often primarily capturing conscious reflections (i.e. what the participant is aware of) but missing habitual, unconscious or socially rooted behaviours (18, 23). While a think-aloud method was used during the second focus group, we did not observe HCPs in clinical practice. Direct observation might have yielded valuable real-world insight into behaviour (23, 24).

### Comparison with existing literature

Prior studies of CVD interventions in primary care have also assessed barriers and facilitators experienced by HCPs. Alageel et al. examined multiple health behaviour change (MBHC) interventions in the NHS Health Check for CVRM in UK primary care, interviewing HCPs to understand the limited effectiveness of these interventions (25). In addition to time constraints and work pressure, barriers identified in this study included patient capabilities, lack of training, and pessimism concerning effectiveness (25). These barriers are acknowledged in the current study design. Supporting our design and results, Molema et al. interviewed Dutch primary care HCPs concerning a combined lifestyle intervention, finding that implementation problems related to the use of health-promoting financial incentives are best examined early on using the Behaviour Change Wheel (26). A 2022 systematic review by Lu et al. examined evidence for clinical decision support interventions on improving CVD outcomes, healthcare processes and implementation barriers (27). The authors reported slow uptake and concluded that adoption is heavily influenced by context and concordant implementation strategies.

Although the relatively novel concept of population-level risk stratification is still emerging in population health management, earlier studies have made efforts in this direction. In a UK study by Chung et al., a model with age- and sex-specific risk thresholds prioritising individuals for CVD risk assessment, resulted in lower screening numbers and more efficient CVD risk assessment (28). However, to our knowledge, our study is the first to use a proactive population health management approach that explores how changes in health care delivery - from reactive to proactive - are perceived by HCPs.

### Implications for practice

A better understanding of identified barriers and facilitators of target behaviours with respect to the PROSPERA programme (step 1 of the Behavioural Change Wheel) could support selection of relevant behaviour change techniques or implementation strategies (17). However, it is essential to consider barriers and facilitators affecting implementation at an early stage. Following the BCW, we considered corresponding behaviour change techniques (BCTs) for identified barriers that could be applied in a future clinical trial (17, 29). For instance, to mitigate panel inaccuracies we could consider alignment with a specific EHR used in most general practices in the greater Leiden and the Hague region, offering practices the most-up-to date information for panel risk stratification and U-Prevent. Barriers relating to omission of the Lifestylecheck or other PROSPERA components could be partially mitigated by restructuring the physical and digital environment, for instance through desktop shortcuts and visible placement of the Lifestylecheck on the HCP’s desk. To address skills, further training in risk communication and pocket cards with instructions could be offered (29, 30). To stimulate engagement, other BCTs could include feedback on behaviour, practical support via the researcher and celebrating successes or providing rewards for the desired target behaviour.

When developing complex interventions, our findings highlight the importance of considering barriers and facilitators informing behaviour change among HCPs at an early stage. We expect this study to be relevant for countries with healthcare systems in which healthcare professionals such as GPs serve a similar gatekeeping role, providing a roadmap for implementation of complex multilevel CVRM interventions in primary care.

## Data Availability

All data relevant to the study are accessible from the author upon reasonable request.

## Provenance and peer review

Not commissioned; externally peer reviewed.

## Ethical approval

The Medical Ethical Review Board of Leiden University Medical Center mandated the review of research not falling under the Dutch Medical Research with Human Subjects Law (nWMO) to individual nWMO committees at Leiden University Medical Center. The nWMO committee reviewed the study proposal and provided a declaration of no objection (non-WMO approval number: 23-3050).

## Data

All data relevant to the study are accessible from the author upon reasonable request.

## Acknowledgements

We thank our partners who contributed to the study: Harteraad, HADOKS, Stichting Langerhans, Stichting Informatievoorziening voor Zorg en Onderzoek (STIZON), Instituut voor Zorgoptimalisatie (INSZO) and ORTEC B.V. We especially thank all general practitioners and practice nurses who participated and generously gave their time and commitment.

